# N-acetyl Cysteine attenuates intrinsic functional connectivity, but not neural alcohol cue reactivity, in treatment-seeking individuals with alcohol use disorder

**DOI:** 10.1101/2023.10.18.23297059

**Authors:** Warren B Logge, Paul S Haber, Tristan P Hurzeler, Ellen E Towers, Kirsten C Morley

## Abstract

N-acetyl cysteine (NAC) is a potential pharmacotherapy for alcohol use disorder (AUD), but it is not known whether it modulates neural activation to alcohol cues or intrinsic functional connectivity. We investigated whether NAC attenuates i) alcohol cue-elicited activation, and ii) intrinsic functional connectivity compared to placebo in patients with AUD. Twenty-three individuals (7 females) with moderate-severe AUD received daily NAC (2400 mg/day, *n* = 9), or a placebo (*n* = 14) for at least 2 weeks. Participants completed a pre-treatment functional magnetic resonance imaging session (T0) and a post-treatment session (T1) comprising a resting-state and visual alcohol cue reactivity task acquisitions. Activation differences between sessions, treatment, and session-by-treatment interaction were assessed. Resting-state functional connectivity examined using 376 node ROI-to-ROIs evaluated whether NAC reduced intrinsic functional connectivity after treatment. There were no differences in alcohol cue reactivity for brain activation or subjective craving between NAC and placebo during treatment or across sessions, or significant interaction. A significant treatment-by-time interaction, with reduced intrinsic connectivity was observed after treatment (T1) for NAC-treated compared to placebo-treated patients in the posterior cingulate node (9, left hemisphere) of the dorsal attentional network and connections to salience, ventral-attentional, somatosensory, and visual-peripheral networks implicated in AUD. NAC reduced intrinsic functional connectivity in patients with moderate-severe AUD after treatment compared to placebo, but did not attenuate alcohol cue-elicited activation. The reduced intrinsic functional connectivity pattern seen may signify reduced external processing of environmental alcohol cues, though no reduced visual cue reactivity associations were evidenced.

## Introduction

Alcohol use disorder (AUD) is a chronic relapsing condition associated with significant burden of disease, yet with limited treatment options (Haber et al., 2021). **S**everal pharmacotherapies exist for the management of AUD. However, these pharmacotherapies have modest effects overall at reducing drinking behaviors (Morley et al., 2021). Further development of effective and novel treatments is urgently required to provide therapeutic options for individuals with AUD.

N-acetyl cysteine (NAC) is proposed to normalize expression of the glutamate transporter (GLT-1) and cystine-glutamate exchange, restoring glutamate homeostasis (Morley et al., 2018). GLT-1 protein levels can be reduced through chronic alcohol intake leading to disruption of glutamatergic input from the prefrontal cortex to reward areas (Hwa et al., 2017). In rodents, compounds that modulate GLT-1 (Sari et al., 2015) and glutamate receptor antagonists (Adams et al., 2010) diminish the reinstatement of both alcohol-seeking behaviors and alcohol intake. Accordingly, glutamatergic agents such as NAC have become candidate pharmacotherapies for the management of addictive behaviors.

NAC has a demonstrated safety profile with signs of efficacy in a range of neuropsychiatric disorders. One meta-analysis of adjunctive NAC demonstrated significant global improvements in mood disorders (Kishi et al., 2020). NAC (2400 mg/day) favorably influenced abstinence and dependence severity in cocaine users, along with improved executive control (Schulte et al., 2018). NAC also reduced daily smoking and depression relative to placebo in people with tobacco use disorder (Prado et al., 2015). One secondary analysis of a 12 week trial of NAC (2400 mg/day) in cannabis use disorder found significant increases in abstinence, reduced weekly drinking and drinking days for NAC versus placebo (Squeglia et al., 2018). Reductions in drinking were similarly observed in a secondary analysis of a trial of NAC for cannabis use disorder in adolescents (Squeglia et al., 2016).

Functional neuroimaging techniques are effective tools to identify biomarkers for AUD pharmacotherapies, can robustly evaluate changes associated with pharmacological treatment (Bach et al., 2022), and highlight potential for novel medications (Grodin and Ray, 2019). However, limited studies have assessed the effects of NAC in modulating cue reactivity in substance use disorder populations, and none have assessed effects in AUD. One study evaluating neural cue reactivity and working memory in regular cocaine-using men found NAC did not modulate cocaine cue-elicited brain activity or elicit marked changes in working memory outcomes after 25 days of treatment (2400 mg/day) versus placebo (Schulte et al., 2019). Additionally, functional brain alterations can also reliably be evaluated using resting-state (or intrinsic) functional connectivity, which measures non task-dependent BOLD fluctuations to characteristic intrinsic brain neuronal activity whilst subjects are awake (Biswal, 2012). Intrinsic functional connectivity is a potentially robust and reliable biomarker of treatment response in alcohol and substance use disorders with particular utility during preliminary testing of novel treatment targets to establish clinical potential (Tolomeo and Yu, 2022, Wilcox et al., 2019). Accordingly, we undertook a double-blind randomized controlled longitudinal pharmaco-fMRI trial of NAC versus placebo in treatment-seeking AUD individuals. We investigated whether N-acetylcysteine (NAC, 2400mg/day) versus placebo modulates neural activation from baseline pre-treatment to after treatment through i) attenuation of alcohol cue reactivity during a visual fMRI task and ii) differences in intrinsic functional connectivity patterns.

## Materials and Method

Participants were randomized to receive placebo, or (2400 mg/day) NAC (1:1 ratio) for 28 days. The study was conducted over a 36-month period (Royal Prince Alfred Hospital) in Australia between 2019 and 2021. The study was approved by the Human Ethics Review Committee of the Sydney Local Health District (X17-0343 & 2019/STE08617). The study involved off-label use of a registered medication in Australia and approval was given under the Clinical Trial Notification (CTN) scheme of the Therapeutics Goods Administration (TGA) (2013/0060). All participants included in this MRI sub study provided written informed consent after commencement of randomization for the main trial.

### Participants

Thirty-four participants were recruited as part of a larger trial investigating the efficacy of NAC in alcohol use disorder (Morley et al., 2023). Inclusion criteria included AUD according to the DSM-V criteria, a desire to reduce or stop drinking, have consumed at least 21 standard drinks per week or 2 heavy drinking days per week (HDD: ≥5 standard drinks/day for men; ≥4 for women) in the month prior to screening, age 18-70, adequate cognition and English language skills to give valid consent and complete research interviews, and willingness to give written informed consent.

Exclusion criteria were pregnancy or lactation, concurrent use of any psychotropic medication other than antidepressants (provided these were taken at stable doses for at least two months); any other substance dependence other than nicotine; clinically unstable medical (e.g. cancer, end stage liver disease) or psychiatric disorder, (e.g. active psychosis or active suicide risk that precluded trial participation; concurrent use of selenium, vitamin D or other antioxidants; and any alcohol pharmacotherapy used within the previous month. One participant presented significant cognitive dysfunction, and one participant had a significant traumatic brain injury identified during baseline scan and their results were not included in the analyses. 12 participants dropped out of the trial before attending their next scan appointment after the baseline and their data excluded from analyses.

### Procedure

Participants underwent a structured interview and medical consultation on day 0 assessing trial eligibility. For the main trial, participants were randomised 1:1 to NAC at 2400 mg/day (2 × 1200 mg capsules twice a day) for 28 days or placebo capsules of identical appearance. Participants completed baseline questionnaires. Imaging sessions were scheduled at baseline prior to first treatment administration (T0) and a follow-up treatment session around 19 days later (SD = 3.73, T1) based on patient availability. Sessions were conducted between 11 am and 4 pm. Participants completed the cue reactivity task approximately 120 minutes after taking their first daily medication capsule.

### Assessments

Consumption in the preceding 28 days before T0 scan day was measured using the Timeline followback interview (TLFB) (Sobell and Sobell, 1992) expressed as the number of Australian standard drinks (10 g ethanol) per drinking day (henceforth TLFB units). For T1, participants completed TLFB reporting days since the T0 scan day. The Penn Alcohol Craving Scale (PACS; (Flannery et al., 1999)) measured subjective craving experienced across the week, and the Alcohol Urge Questionnaire (AUQ; (Bohn et al., 1995)) was completed pre-and post scan assessing changes in state craving and urges to drink, with higher scores indicating greater craving, The Alcohol Dependence Scale (ADS) (Skinner and Allen, 1982) measured participants’ AUD severity, with higher scores indicating greater severity and only completed at T0.

### fMRI Cue Reactivity

A well-established visual cue reactivity task (Grüsser et al., 2004) was used to measure alcohol cue-elicited brain activity. Stimuli comprised 15 alcohol-related pictures depicting types of alcohol and drinking situations (Grüsser et al., 2004, Wrase et al., 2002), and 30 control images: 15 affective neutral pictures from the International Affective Picture System (Lang et al., 1997) matched for colour and complexity, and 15 scrambled versions of the scrambled alcohol pictures controlling for potential activity related to novelty of neutral images. Images were presented for 6.6 s in blocks of three images of the same type, totalling five blocks per type (alcohol, affective neutral). Stimuli and block order were randomised across subjects, and blocks of the same image type did not follow consecutively. Each condition block was preceded by a fixation cross presented for 10 s modelled as a regressor of no interest. The total task duration was 520 s. Participants were debriefed after test session to address potential continued craving elicited during scanning.

### MRI Data Acquisition

MRI data were acquired on a 3-Tesla GE Discovery (GE Healthcare, Milwaukee, Wisconsin, United States) using a 32-channel head coil. A T1-weighted (1-mm^3 voxel resolution) structural scan was acquired for each subject for screening and registration (TR: 7200 ms, TE 2.7 ms, 176 sagittal slices, 1 mm thick, no gap, 256 × 256 × 256 matrix). For BOLD acquisitions, we acquired echoplanar image (EPI) volumes (cue reactivity: n = 183; resting state: n = 200) comprising 39 axial slices in an ascending interleaved fashion with a voxel resolution of 1.88 × 1.88 × 2 mm (TR: 3000 ms, TE 30 ms, FA 90 degrees, FOV 240 mm, matrix 128 × 128, acceleration factor 2, slice gap: 1 mm). Participants’ heads were fixated with foam pads to minimise head movement. For fMRI resting-state acquisition participants were instructed to keep their eyes closed and to focus on nothing in particular.

### Image Processing

A detailed overview of imaging processing pipeline is provided in the Supplementary Material, with a summary provided here. Anatomical and functional preprocessing was completed using FMRIPrep ((Esteban et al., 2020, Esteban et al., 2019), RRID:SCR_016216), with the two session T1-weighted structural images corrected for intensity non-uniformity, skull-stripped, and then used for reconstruction of the brain surfaces. Volume-based structural images were segmented into white matter, grey matter, and cerebrospinal fluid, and then spatially normalised into MNI space.

Functional MRI data for the two sessions per subject and per acquisition (cue reactivity, resting-state) were pre-processed with susceptibility distortion correction using a B0-nonuniformity map (i.e., fieldmap) based on two EPI references in opposing phase-encoding directions, and slice-timing correction was completed. Images underwent motion correction, co-registration to structural data, normalisation to MNI space and projection to cortical surface. Functional timeseries were then resampled to FreeSurfer’s (FreeSurfer 6.0.1, surfer.nmr.mgh.harvard.edu) fsaverage space. Cue reactivity data were post-processed within SPM, with functional resampled images smoothed with a Gaussian kernel of 8 mm full-width half maximum (FWHM) to improve sensitivity for group analysis. Resting-state data were further post-processed using eXtensible Connectivity Pipelines (XCP-D) (Ciric et al., 2017, Satterthwaite et al., 2013). Volumes with framewise-displacement (FD) greater than 0.4 were flagged as outliers and excluded. A total of 36 nuisance regressors were regressed out of the BOLD data using the ‘36P’ strategy (Ciric et al., 2017, Satterthwaite et al., 2013). Residual timeseries were then band-pass filtered (0.01-0.08 Hz) and spatially smoothed with a kernel size of 6mm (FWHM). Fisher’s r-to-z transformation was applied to the Pearson correlation coefficients per cell of the resultant matrices of the ROI-to-ROI connectivity matrix using the Schaefer 17-network 400 parcel atlas (Schaefer et al., 2018) for each subject outputted from the XCP-D processing. Of the 400 parcels from the 17 functional networks, 23 parcels contained missing data and were excluded from second-level analyses (See supplementary Material for information), leaving a 377×377 symmetrical functional connectivity matrix per participant per session (46 matrices total).

### Statistical analysis

Differences in treatment group sample demographics, clinical and drinking measures, and PACS craving were assessed by two-sample t-tests or Mann-Whitney U tests, where appropriate. We used a linear mixed effects model (LMM) with a random intercept of participant to assess any group differences between pre- and post-scan session AUQ craving compared across T0 and T1 sessions.

fMRI cue reactivity analyses were conducted in SPM12 at two levels. Two conditions were modelled at the first level (subject-specific): alcohol-related (Alcohol) cues; and both control cues combined in a single condition (Control) to control for both novelty and visual complexity of the stimuli, modelled as a box-car function convolved with the canonical haemodynamic response.

Motion correction parameters (six regressors) also modelled at the first level as regressors of no interest. The fixation cross was left as an implicit baseline.

Regions of interest (ROIs) were selected based on areas correlated with alcohol cue reactivity identified by a recent meta-analysis evaluating pharmacotherapy studies in AUD (Zeng et al., 2021). Five ROIs were used: the left and right caudate, the left and right dorsolateral prefrontal cortex (DLPFC), and bilateral ventromedial prefrontal cortex (VMPFC). The caudate was defined here as caudate body from the Harvard-Oxford subcortical probability atlas (http://www.cma.mgh.harvard.edu/fsl_atlas.html). The DLPFC and VMPFC were probabilistic maps defined using the Brainmap database (Fox and Lancaster, 2002) binarized with a threshold of ≥.90. We used the MarsBar toolbox (Brett et al., 2002) to extract the ROI unweighted beta estimates (*β*) for the baseline and treatment scans which were then exported into R software (Version 4.0.3) for further analyses using LMMs to assess group treatment differences in ROI activity while accounting for other variables. To optimally balance Type I and Type II error associated with multiple comparisons across ROIs, we accounted for the correlation between the dependent variables of the five probabilistic ROI using the Simple Interactive Statistical Analysis Bonferonni tool (http://www.quantitativeskills.com/sisa/calculations/bonfer.htm). We applied a method outlined in Li et al. (2014), whereby the resultant Bonferroni correction is less stringent as it does not assume the variables are obtained from independent subgroups, as would occur in a standard and more conservative Bonferroni correction. The 5 ROI betas had a mean correlation coefficient of *r* = .066 (number of tests *k* = 5), resulting in an equivalent corrected alpha of .029 which would be equivalent to a corrected P <.05.

As difference contrast comparisons of Alcohol > Control have low reliability for repeated scans (Bach et al., 2022) the individual contrast images for the Alcohol and Control image conditions were used for second-level models. LMMs were conducted comparing ALC or CON image type across sessions (T0, T1), treatment, and a session-by-treatment interaction. Patient age was added to the models to account for differences in brain activation associated with age and controlling for additional years of harmful drinking seen in older drinkers. Antidepressant use was added as a covariate due to the potential augmentative effects of NAC associated with antidepressant use and mixed evidence regarding improved outcomes when antidepressants and NAC are administered concurrently, possibly due to reductions in oxidative stress (Costa-Campos et al., 2013). Presence of alcohol-related liver disease (ArLD) was added due to its effects on cortical concentrations of GABA (Morley et al., 2020) and potential impacts on cue reactivity related to symptoms (Logge et al., 2019).

ANCOVAs were conducted for T0 and T1 respectively for the ALC > CON contrast to evaluate ROI beta activity at baseline and after treatment and are presented in Supplementary Material. Age was included as a covariate in the analyses for both sessions. The T0 ANCOVA included TLFB drinks per drinking day prior to enrolment, and respective interactions, to evaluate the effect of baseline drinking on cue reactivity. The T1 ANCOVA included TLFB drinks per drinking day as a main effect, and number of treatment days as an additional covariate along with respective interactions to control for the time of T1 scan based on patient availability.

We performed additional exploratory whole brain analyses to examine brain activity not evaluated by the a priori ROIs using the ‘Multivariate and Repeated Measures for Neuroimaging’ (MRM) toolbox (version 1.0, (McFarquhar et al., 2016)) a multivariate and permutation approach which allows advanced statistical modelling of repeated measures mixed effects designs using a multivariate form of the general linear model employed within Matlab. First level contrasts (ALC, CON) were entered into respective second-level full factorial whole-brain general linear models (GLM) comparing NAC and placebo using two sample *t*-tests to assess group differences in alcohol and control cue reactivity across the two sessions (T0 and T1). Statistical thresholds were set using permutation-based inference, with 5000 permutations conducted to appropriately account for the within and between-subjects’ variance in the model while assessing the betas for the two conditions. Whole-brain analyses were corrected with a family-wise error cluster level inference (pFWEc) used at < 0.05 with a cluster-forming height threshold of p = .001, in accordance with recommendations by Eklund et al. (2016). MNI coordinates are reported in x, y, z dimensions, with regions identified using the SPM Wake Forest University (WFU) Pickatlas toolbox (http://www.fmri.wfubmc.edu/cms/software, version 3.0.5). Whole-brain results are reported in the Supplementary Material.

Resting state data were analysed using CONN toolbox (Whitfield-Gabrieli and Nieto-Castanon, 2012) version 22a (Nieto-Castanon and Whitfield-Gabrieli, 2021). The 46 377×377 ROI-to-ROI z score correlation matrices were entered into a second-level GLM model with within-subject contrast session (T0 < T1 [i.e., -1 1]) evaluating post-treatment effects and between-subjects treatment (NAC > placebo [i.e., 1 -1]) evaluating the effect of NAC. Age, antidepressant use, and ArLD were added as covariates. A parametric multivariate statistics approach was conducted to target the contributions of the individual ROIs. An FDR-corrected ROI-level p-value (MVPA omnibus test) was used with a threshold of PFDR <.05 (Benjamini and Hochberg, 1995) with a connection (height) threshold of p <.01 (uncorrected) to examine the individual connections between the ROIs.

## Results

### Sample Characteristics and Drinking Outcomes

Table 1 summarises the demographic and clinical characteristics of the sample and drinking outcomes. There were no significant differences between placebo and NAC groups for age, gender, years of education, or unemployment. Both groups demonstrated similar levels of TLFB drinks per drinking day prior to treatment, number of years since alcohol-related problems began, and levels of alcoholic liver disease, tobacco use, and anti-depressant use (*p*’s > .269). There were no group differences in severity of alcohol dependence measured by ADS (p > 386).

**Table 1.** Sample Characteristics and Subjective Craving Scores with Tests of Group Differences.

|  | Placebo n = 14 | NAC n = 9 |
| --- | --- | --- |
| *Demographics* |  |  |
| Age | 48.1 ± 11.57 | 51.67 ± 12.72 |
| Gender, % F | 21 | 44 |
| Education, y | 14.36 ± 2.38 | 14.64 ± 3.04 |
| Unemployed, % | 18.18 | 16.7 |
| *Clinical Characteristics* |  |  |
| Baseline drinks per drinking day <sup>a</sup> | 16.96 ± 7.86 | 11.97 ± 10.27 |
| Baseline drinking days per week <sup>a</sup> | 3.1 ± 2.95 | 2.7 ± 2.87 |
| Years since alcohol-related problems began | 20 ± 13.42 | 16.27 ± 10.51 |
| ADS score | 20.35 ± 10.73 | 24.22 ± 9.82 |
| Alcoholic liver disease, % | 36 | 22 |
| Tobacco use, % | 72.72 | 90.9 |
| Anti-depressant use, % | 43 | 56 |
| Treatment days between scans | 15 ± 4.34 | 14 ± 2.7 |
| *Subjective Craving* |  |  |
| PACS Craving Baseline | 14.77 ± 8.95 | 17.67 ± 7.97 |
| PACS Craving during treatment | 14.62 ± 7.92 | 15.71 ± 8.8 |
| AUQ |  |  |
| T0: pre-scan | 18.21 ± 10.03 | 15.11 ± 10.79 |
| T0: post-scan | 19 ± 9.25 | 15.22 ± 11.95 |
| T1: pre-scan | 18.54 ± 10.56 | 15.11 ± 10.86 |
| T1: post-scan | 20.54 ± 9.72 | 12.9 ± 7.8 |

*Note.* Data represent $M \pm SDs$ , unless otherwise noted. NAC = N-Acetyl cysteine; ADS = Alcohol Dependence Severity Scale; PACS = Penn Alcohol Craving Scale; AUQ = Alcohol Urge Questionnaire.
<sup>a</sup> During the 28 days preceding the first day of the study, based on the Time-Line Follow-Back method.

### Subjective Craving

Craving was reported both for the previous week using PACS and pre- and post-scan across both sessions with mean AUQ scores presented in Table 1. No differences in PACS subjective craving were seen between groups, between sessions, or interactions (p’s > .731). AUQ scores during sessions showed no differences according to treatment group, pre- versus post-scan, or between sessions, or interactions (p’s > 104) . LMMs of AUQ scores are presented in Supplementary Material. The LMM for T0 assessing baseline and T1 only assessing changes in craving after treatment found no main treatment or pre- versus post-scan differences in AUQ score. However, there a significant main effect of age for the T1 model, with older participants reporting less overall craving during treatment (β **=** -0.33, p = .038).

### Alcohol Cue Reactivity

The contrasts for the separate image types (ALC, CON) are used to evaluate treatment across sessions. The LMM model tables for these ALC and CON contrasts for 5 ROIs are presented in Table 2 and Table 3. The ALC v CON contrast tables at T0, T1, and across sessions are further presented in the Supplementary material for reference.

**Table 2.** Linear Mixed Effects model for ALC contrast across 5 ROIs.

|  | Bl VMPFC |  |  | L Caudate |  |  | R Caudate |  |  | L DLPFC |  |  | R DLPFC |  |  |
| --- | --- | --- | --- | --- | --- | --- | --- | --- | --- | --- | --- | --- | --- | --- | --- |
| Predictors | $\beta$ | CI | p | $\beta$ | CI | p | $\beta$ | CI | p | $\beta$ | CI | p | $\beta$ | CI | p |
| (Intercept) | 0.49 | -1.06 – 2.03 | .537 | -0.13 | -1.56 – 1.30 | .855 | -0.03 | -1.27 – 1.21 | .964 | -0.38 | -1.17 – 0.41 | .343 | -0.37 | -1.31 – 0.57 | .443 |
| Session | -0.35 | -0.80 – 0.09 | .122 | -0.51 | -1.27 – 0.25 | .192 | -0.58 | -1.24 – 0.08 | .084 | -0.29 | -0.64 – 0.06 | .109 | -0.54 | -1.04 – -0.04 | .034 |
| Treatment | -0.18 | -0.99 – 0.63 | .665 | -0.79 | -1.68 – 0.11 | .085 | -0.76 | -1.53 – 0.02 | .055 | 0.01 | -0.45 – 0.47 | .969 | 0.13 | -0.46 – 0.72 | .676 |
| Age | 0 | -0.03 – 0.03 | .816 | 0.01 | -0.02 – 0.04 | .455 | 0.01 | -0.02 – 0.03 | .55 | 0.01 | -0.01 – 0.02 | .23 | 0.01 | -0.01 – 0.03 | .256 |
| Antidepressant Use | -0.08 | -0.82 – 0.66 | .838 | 0.52 | -0.14 – 1.19 | .124 | 0.49 | -0.09 – 1.07 | .098 | -0.02 | -0.39 – 0.35 | .912 | -0.19 | -0.63 – 0.25 | .401 |
| Alcoholic Liver Disease | -0.14 | -0.89 – 0.62 | .725 | 0.22 | -0.46 – 0.90 | .531 | 0.03 | -0.56 – 0.62 | .915 | 0.19 | -0.19 – 0.57 | .335 | 0.36 | -0.09 – 0.81 | .113 |
| Session * Treatment | 0.05 | -0.66 – 0.76 | .889 | 0.07 | -1.14 – 1.29 | .905 | 0.36 | -0.70 – 1.41 | .506 | 0.15 | -0.41 – 0.72 | .598 | 0.05 | -0.75 – 0.85 | .907 |
| **Random Effects** |  |  |  |  |  |  |  |  |  |  |  |  |  |  |  |
| $\sigma^2$ | 0.36 | | | 1.05 | | | 0.79 | | | 0.23 | | | 0.46 | | |
| $\tau_{00}$ | 0.46 ID | | | 0.00 ID | | | 0.00 ID | | | 0.05 ID | | | 0.00 ID | | |
| ICC | 0.56 |  |  |  |  |  |  |  |  | 0.18 |  |  |  |  |  |
| N | 23 ID |  |  | 23 ID |  |  | 23 ID |  |  | 23 ID |  |  | 23 ID |  |  |
| Observations | 46 |  |  | 46 |  |  | 46 |  |  | 46 |  |  | 46 |  |  |
| Marginal R<sup>2</sup> | 0.049 |  |  | 0.183 |  |  | 0.166 |  |  | 0.106 |  |  | 0.197 |  |  |

P-threshold < .029 (Bonferroni's adjustment corrected)
Note. Reference category for predictor shown in brackets. $\sigma^2$ = random effects variance; $\tau_{00}$ = random intercept variance ICC = intra-class correlation coefficient; ID = individual participants (random factor); Bl = bilateral, L = left, R = right, DLPFC = dorsolateral prefrontal cortex.

**Table 3.** Linear Mixed Effects model for CON contrast across 5 ROIs.

|  | BI VMPFC |  |  | L Caudate |  |  | R Caudate |  |  | L DLPFC |  |  | R DLPFC |  |  |
| --- | --- | --- | --- | --- | --- | --- | --- | --- | --- | --- | --- | --- | --- | --- | --- |
| Predictors | $\beta$ | CI | p | $\beta$ | CI | p | $\beta$ | CI | p | $\beta$ | CI | p | $\beta$ | CI | p |
| (Intercept) | 0.27 | -1.24 – 1.77 | .729 | -0.01 | -1.25 – 1.23 | .986 | 0.03 | -1.04 – 1.11 | .951 | -0.34 | -1.05 – 0.37 | .346 | -0.26 | -1.09 – 0.57 | .539 |
| Session | -0.22 | -0.66 – 0.23 | .34 | -0.51 | -1.16 – 0.15 | .131 | -0.57 | -1.14 – 0.00 | .051 | -0.22 | -0.53 – 0.09 | .168 | -0.42 | -0.86 – 0.02 | .062 |
| Treatment | -0.12 | -0.91 – 0.68 | .774 | -0.88 | -1.66 – -0.11 | **.025** | -0.83 | -1.50 – -0.16 | **.015** | -0.01 | -0.42 – 0.41 | .976 | 0.12 | -0.40 – 0.64 | .642 |
| Age | 0 | -0.03 – 0.03 | .836 | 0.01 | -0.01 – 0.03 | .45 | 0.01 | -0.01 – 0.03 | .548 | 0.01 | -0.00 – 0.02 | .176 | 0.01 | -0.01 – 0.02 | .251 |
| Antidepressant Use | 0.06 | -0.66 – 0.78 | .878 | 0.62 | 0.04 – 1.19 | .037 | 0.56 | 0.06 – 1.06 | .03 | 0.03 | -0.30 – 0.37 | .853 | -0.14 | -0.53 – 0.24 | .472 |
| Alcoholic Liver Disease | -0.24 | -0.98 – 0.49 | .514 | 0.08 | -0.51 – 0.67 | .799 | -0.06 | -0.57 – 0.46 | .829 | 0.05 | -0.29 – 0.39 | .783 | 0.2 | -0.20 – 0.59 | .325 |
| Session * Treatment | -0.13 | -0.85 – 0.58 | .711 | 0.03 | -1.02 – 1.08 | .954 | 0.38 | -0.54 – 1.29 | .42 | 0.03 | -0.47 – 0.53 | .916 | -0.03 | -0.73 – 0.68 | .9 |
| **Random Effects** |  |  |  |  |  |  |  |  |  |  |  |  |  |  |  |
| $\sigma^2$ | 0.36 | | | 0.79 | | | 0.59 | | | 0.18 | | | 0.35 | | |
| $\tau_{00}$ | 0.43 ID | | | 0.00 ID | | | 0.00 ID | | | 0.04 ID | | | 0.00 ID | | |
| ICC | 0.54 |  |  |  |  |  |  |  |  | 0.2 |  |  |  |  |  |
| N | 23 ID |  |  | 23 ID |  |  | 23 ID |  |  | 23 ID |  |  | 23 ID |  |  |
| Observations | 46 |  |  | 46 |  |  | 46 |  |  | 46 |  |  | 46 |  |  |
| Marginal R<sup>2</sup> | 0.045 |  |  | 0.254 |  |  | 0.225 |  |  | 0.093 |  |  | 0.164 |  |  |

P-threshold < .029 (Bonferroni's adjustment corrected)
Note. Reference category for predictor shown in brackets. $\sigma^2$ = random effects variance; $\tau_{00}$ = random intercept variance ICC = intra-class correlation coefficient; ID = individual participants (random factor) ; BI = bilateral, L = left, R = right, DLPFC = dorsolateral prefrontal cortex.

When evaluating reactivity to ALC cues across T0 and T1 sessions, there were no significant effects seen for treatment group, session, or two-way interaction seen for any of the 5 ROIs. This was similar for CON cues across T0 and T1 sessions, with no significant main effects or interactions seen for the 5 ROIs.

#### Treatment Effects

For ALC only, no main effects of treatment were seen for the 5 ROIs averaging across the samples. For CON only, there was a significant main effect of treatment seen in the left caudate (p = .022) and right caudate (p = .015), with NAC-treated participants showing decreased control cue-elicited activation than placebo-treated participants across sessions overall.

#### Covariates

For ALC only, there was a main effect of antidepressants seen for the left caudate (p = .020) and right caudate (p = .019), with patients not taking antidepressants, demonstrating lower alcohol cue-elicited activation than those taking antidepressants, controlling for age and ArLD. However, similar activations were seen for CON only with a main effect of lower alcohol cue-elicited activation in patients not taking antidepressants in these two ROIs (left caudate p = .021, right caudate p = .022), suggesting a generalized response to cues. No other main effects were seen for the covariates.

### Resting state/intrinsic functional connectivity

There was a significant treatment-by-time interaction seen with reduced functional connectivity patterns observed in the NAC participants compared to placebo after treatment in one ROI seed region, posterior cingulate 9 (MVPA PFDR = .01) in the dorsal attentional (B) network of the Schaefer 400 atlas parcellation. The connectivity target regions included those in the left and right somatosensory networks, left temporal-parietal network, bilateral visual-central and visual-peripheral regions, and salience ventral attentional networks. Fig. 1 presents these connections, with connectivity values reported in Supplementary Material. No evidence of other significant connectivity patterns was seen at either the session or treatment levels, or interaction.

**Fig 1.**
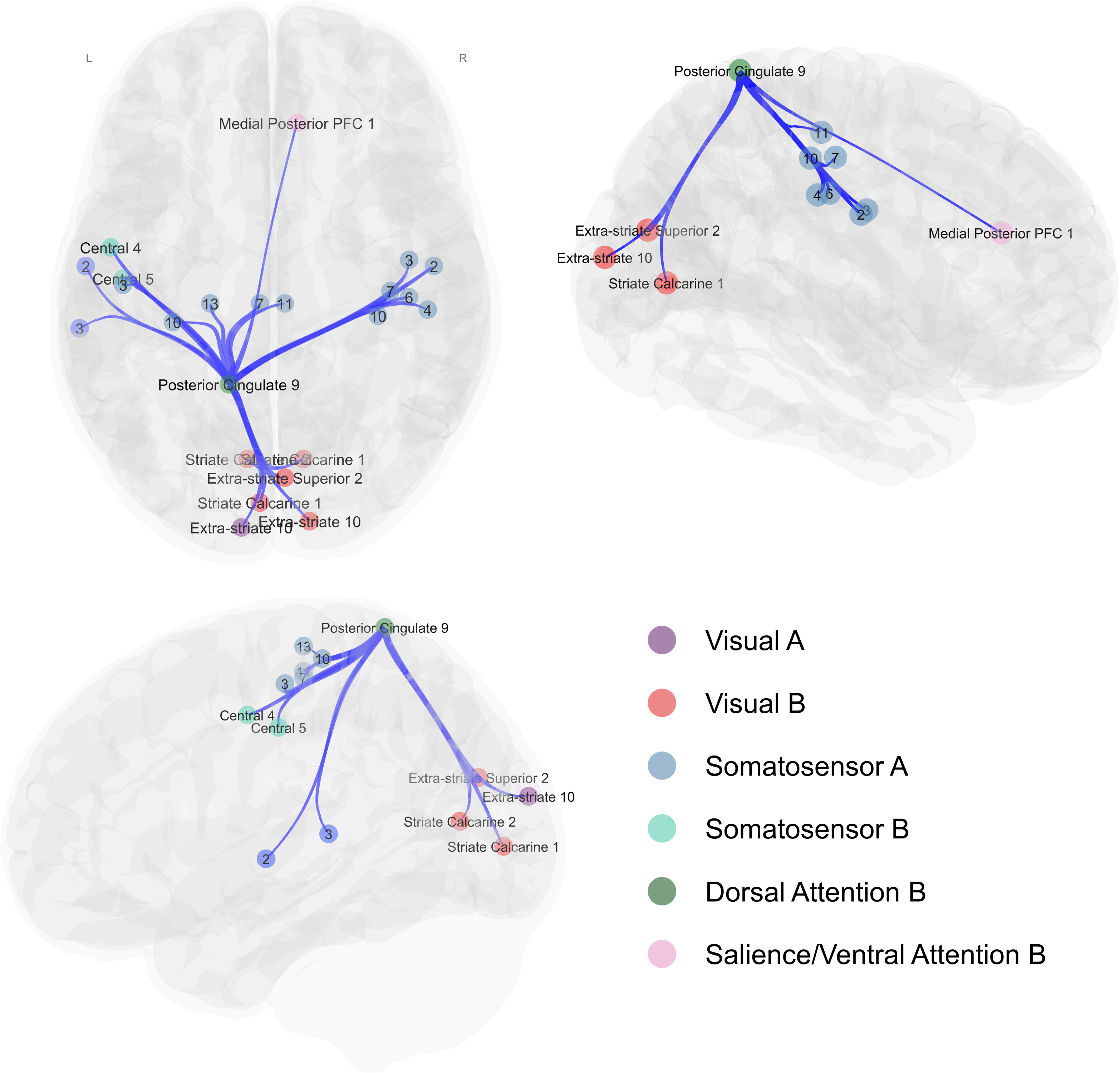
Axial (top-left panel), right sagittal (right panel) and left sagittal (bottom-left panel) views of the significant ROI-to-ROI treatment-by-session interaction. Spheres represent Schaefer-400 parcellation nodes (regions of interest) for the significant connections between the posterior cingulate (dark green), with lines representing connections between nodes that were significant at a nominal, uncorrected p-value threshold of 0.01. Networks are labelled in the figure are color-coded according to Yeo et. al (2011) networks.

## Discussion

This is the first study examining the effects of NAC on fMRI alcohol cue reactivity and intrinsic functional connectivity in patients with AUD. We observed that patients with AUD demonstrated reduced intrinsic functional connectivity after NAC treatment compared to placebo-treated patients. However, no evidence of NAC treatment effects was seen for alcohol cue-elicited reactivity, either across sessions, or a session-by-treatment interaction, indicating specific modulation in functional brain activation after NAC treatment.

This reduced intrinsic connectivity after NAC treatment was seen in a PCC node associated with the dorsal attentional network with projections to prefrontal salience attentional network node, multiple somatosensory network nodes, and visual network nodes. The PCC is considered a connective neural hub region with the highest global connectivity of any brain region, demonstrating modulated activation patterns during rest and active states (Pfefferbaum et al., 2011, Hagmann et al., 2008). The PCC is a well-established region within the default mode network, but also plays a role in active states involved in processing of external salience of cues, known as exteroceptive processing (Fransson, 2005). Relevant to AUD, this corresponds to responding to processing alcohol-related cues in the environment, with increased activation in the PCC and neighboring precuneus to alcohol cues (Tapert et al., 2003) and PCC activation to alcohol cues the most distinctive brain region in distinguishing AUD and healthy controls (Schacht et al., 2013). The PCC’s active and ‘passive’ roles appear integral in AUD and drinking outcomes. While blunted cingulate cortex connectivity during stress and alcohol cues was implicated in increased risk of relapse in AUD patients compared to healthy controls (Zakiniaeiz et al., 2017), increased connectivity in both AUD and substance use samples characterizes over-awareness or hypervigilance to these cues and is thus disadvantageous (DeWitt et al., 2015). Correspondingly, the reduced connectivity in our study may relate to decreased hypervigilance and sensitization of awareness to external cues. Notably, while the PCC was not an a priori ROI for our cue reactivity analyses, we nevertheless observed alcohol cue reactivity in a cluster in the PCC and precuneus (see Supplementary Material), indicating the PCC is active in these patients with AUD associated with external cue salience processing, though no evidence of treatment effects on alcohol cue reactivity were seen.

Our study is the first to demonstrate the modulatory effect NAC on functional connectivity in AUD. The only other study that evaluated the effects of NAC on intrinsic functional connectivity in substance use investigated smokers (Froeliger et al., 2015) and found an opposing pattern of increased functional connectivity in NAC-treated smokers between the a priori seed nucleus accumbens and fronto-striatal DMN target regions, including the precuneus. This increased connectivity was associated with reduced craving and increased feelings of positive affect (Froeliger et al., 2015), which does not correspond to findings from Huang et al. (2014) that intrinsic functional connectivity in DMN regions, including the PCC, is increased in smokers compared to nonsmokers largely associated with nicotine withdrawal, and the strength of connectivity corresponded with withdrawal-induced increased craving. Moreover, Froeliger et al.’s (2015) study was limited in the cross-sectional approach whereby the 3-day medication paradigm did not allow for a baseline scan to demonstrate initial fronto-striatal-DMN connectivity dysregulation. A strength of our current study is our pre/post-treatment acquisition approach which may explain the our findings of reduced modulation pattern observed post-treatment. Additionally, our whole brain data-driven parcellation approach facilitates the identification of connections that would not be highlighted using a seed-based approach (Lv et al., 2018). Taken together, our results indicate that assessing intrinsic functional connectivity may be an informative approach to investigate the neuropharmacological effect of NAC, and warrants further research in alcohol and substance use disorder samples.

The mechanism by which NAC reduces intrinsic functional connectivity is unclear. One explanation is the normalization of glutamate functioning through activation of inhibitory metatropic glutamatergic receptors, coupled with restoration of GLT-1 which is downregulated after chronic heavy alcohol use (Hwa et al., 2017). Much of the preclinical research focuses on the effects of NAC on striatal regions, such as the nucleus accumbens, to reduce craving (Baker et al., 2003a, Krista et al., 2003, Kalivas et al., 2005, Baker et al., 2003b). However, we notably did not see changes in subjective craving across our trial, though this may be partly explained by overall low craving scores at baseline. Interestingly, NAC-treated patients with AUD showed significantly reduced alcohol consumption in the early stages of the primary trial (i.e., after 7 days) (Morley et al., 2023) but not after four weeks, suggesting NAC may be more effective at the beginning of treatment. This is pertinent to the current findings as the participants’ treatment scan occurred after three weeks of treatment, and our low reported craving scores could be related to minimal differences in between NAC and placebo groups overall drinking toward trial end (Morley et al., 2023).

We did not observe any treatment differences between NAC and placebo on alcohol cue-elicited reactivity across the sample or across sessions. The only other study examining NAC’s effects on neural cue reactivity in cocaine-using men similarly found no modulation of cocaine cue-elicited brain activity after 25 days of NAC treatment versus placebo (Schulte et al., 2019). Notably, minimal baseline cue reactivity in the overall sample before treatment commencement was evidenced, and in our study we similarly observed an overall lack of baseline cue reactivity at T0. As our participants were drinking significantly at baseline and drinking cessation was not required prior to trial commencement plus low overall drinking in the trial, the lack of alcohol cue reactivity may be related to continued alcohol consumption for the majority of participants. Moreover, the marked reduction in drinking for NAC-treated patients that was apparent earlier in the primary trial findings (Morley et al., 2023) as addressed earlier may not have been discernable at T1 scan session which was completed around 3 weeks of treatment.. Interestingly, we did observe some treatment effects associated with control cue reactivity across the sample when assessing CON>CON across the sessions, with reduced reactivity in the NAC group in the left and right caudate. However, with reduced reactivity across the sessions overall, it is likely that with the limited sample size we did not have the power to delineate any changes across sessions of NAC reducing overall reactivity to visual cues in the dorsal striatum. Thus, a larger sample may be required to determine any treatment effect of this activation pattern to visual cues overall.

Our study is the first to demonstrate that NAC treatment attenuated intrinsic functional connectivity in patients with AUD associated with the PCC and key connections in somatomotor, visual, and attentional salience networks. This reduced connectivity pattern may modulate external processing of environmental alcohol cues, yet NAC did not affect visual alcohol cue reactivity, indicating specific, rather than generalized, functional brain activation effects in individuals with moderate-severe AUD. Considering that NAC reduces consumption in the early stages of treatment, future research that comprehensively evaluates the association of intrinsic functional connectivity with drinking outcomes may elucidate the neurobiological effects of NAC on modulating intrinsic connectivity after NAC treatment, and help determine how NAC can be best utilized as a pharmacotherapy for AUD.

## Author Contributions

Warren Logge was involved in the conception and design, acquisition, analysis, interpretation of the data, and original drafting. Paul Haber was involved in the conception, the interpretation of the data, and original drafting. Ellen Towers was involved in the data acquisition and analysis. Tristan Hurzeler was involved in the data acquisition and analysis. Kirsten Morley was involved in the conception and design, interpretation of the data, and original drafting. All authors give final approval for the publication and agree to be accountable for all aspects of this work.

## Data Availability

All data produced in the present study are available upon reasonable request to the authors.

## References

1. Adams C, Short J, Lawrence A (2010) Cue-conditioned alcohol seeking in rats following abstinence: involvement of metabotropic glutamate 5 receptors. Br. J. Pharmacol. 159:534–542.

2. Bach P, Reinhard I, Koopmann A, Bumb JM, Sommer WH, Vollstädt-Klein S, Kiefer F (2022) Test– retest reliability of neural alcohol cue-reactivity: Is there light at the end of the magnetic resonance imaging tube? Addict. Biol. 27:e13069.

3. Baker DA, McFarland K, Lake RW, Shen H, Tang X-C, Toda S, Kalivas PW (2003a) Neuroadaptations in cystine-glutamate exchange underlie cocaine relapse. Nat. Neurosci. 6:743–749.

4. Baker DA, McFarland K, Lake RW, Shen HUI, Toda S, Kalivas PW (2003b) N-Acetyl Cysteine-Induced Blockade of Cocaine-Induced Reinstatement. Ann. N. Y. Acad. Sci. 1003:349–351.

5. Benjamini Y, Hochberg Y (1995) Controlling the False Discovery Rate: A Practical and Powerful Approach to Multiple Testing. Journal of the Royal Statistical Society: Series B (Methodological) 57:289–300.

6. Biswal BB (2012) Resting state fMRI: A personal history. Neuroimage 62:938–944.

7. Bohn MJ, Krahn DD, Staehler BA (1995) Development and Initial Validation of a Measure of Drinking Urges in Abstinent Alcoholics. Alcohol: Clinical and Experimental Research 19:600–606.

8. Brett M, Anton J-L, Valabregue R, Poline J-B (2002) Region of interest analysis using an SPM toolbox.1.

9. Ciric R, Wolf DH, Power JD, Roalf DR, Baum GL, Ruparel K, Shinohara RT, Elliott MA, Eickhoff SB, Davatzikos C, Gur RC, Gur RE, Bassett DS, Satterthwaite TD (2017) Benchmarking of participant-level confound regression strategies for the control of motion artifact in studies of functional connectivity. Neuroimage 154:174–187.

10. Costa-Campos L, Herrmann AP, Pilz LK, Michels M, Noetzold G, Elisabetsky E (2013) Interactive effects of N-acetylcysteine and antidepressants. Prog. Neuropsychopharmacol. Biol. Psychiatry 44:125–130.

11. DeWitt SJ, Ketcherside A, McQueeny TM, Dunlop JP, Filbey FM (2015) The hyper-sentient addict: an exteroception model of addiction. The American Journal of Drug and Alcohol Abuse 41:374–381.

12. Eklund A, Nichols TE, Knutsson H (2016) Cluster failure: Why fMRI inferences for spatial extent have inflated false-positive rates. Proceedings of the National Academy of Sciences 113:7900–7905.

13. Esteban O, Ciric R, Finc K, Blair RW, Markiewicz CJ, Moodie CA, Kent JD, Goncalves M, DuPre E, Gomez DEP, Ye Z, Salo T, Valabregue R, Amlien IK, Liem F, Jacoby N, Stojić H, Cieslak M, Urchs S, Halchenko YO, Ghosh SS, De La Vega A, Yarkoni T, Wright J, Thompson WH, Poldrack RA, Gorgolewski KJ (2020) Analysis of task-based functional MRI data preprocessed with fMRIPrep. Nat. Protoc. 15:2186–2202.

14. Esteban O, Markiewicz CJ, Blair RW, Moodie CA, Isik AI, Erramuzpe A, Kent JD, Goncalves M, DuPre E, Snyder M, Oya H, Ghosh SS, Wright J, Durnez J, Poldrack RA, Gorgolewski KJ (2019) fMRIPrep: a robust preprocessing pipeline for functional MRI. Nature Methods 16:111–116.

15. Flannery BA, Volpicelli JR, Pettinati HM (1999) Psychometric Properties of the Penn Alcohol Craving Scale. Alcohol: Clinical and Experimental Research 23:1289–1295.

16. Fox PT, Lancaster JL (2002) Opinion: Mapping context and content: the BrainMap model. Nat. Rev. Neurosci. 3:319–321.

17. Fransson P (2005) Spontaneous low-frequency BOLD signal fluctuations: An fMRI investigation of the resting-state default mode of brain function hypothesis. Hum. Brain Mapp. 26:15–29.

18. Froeliger B, McConnell PA, Stankeviciute N, McClure EA, Kalivas PW, Gray KM (2015) The effects of N-Acetylcysteine on frontostriatal resting-state functional connectivity, withdrawal symptoms and smoking abstinence: A double-blind, placebo-controlled fMRI pilot study. Drug Alcohol Depend. 156:234–242.

19. Grodin EN, Ray LA (2019) The Use of Functional Magnetic Resonance Imaging to Test Pharmacotherapies for Alcohol Use Disorder: A Systematic Review. Alcohol: Clinical and Experimental Research 43:2038–2056.

20. Grüsser SM, Wrase J, Klein S, Hermann D, Smolka MN, Ruf M, Weber-Fahr W, Flor H, Mann K, Braus DF, Heinz A (2004) Cue-induced activation of the striatum and medial prefrontal cortex is associated with subsequent relapse in abstinent alcoholics. Psychopharmacology (Berl.) 175:296–302.

21. Haber PS, Riordan BC, Winter DT, Barrett L, Saunders J, Hides L, Gullo M, Manning V, Day CA, Bonomo Y, Burns L, Assan R, Curry K, Mooney-Somers J, Demirkol A, Monds L, McDonough M, Baillie AJ, Clark P, Ritter A, Quinn C, Cunningham J, Lintzeris N, Rombouts S, Savic M, Norman A, Reid S, Hutchinson D, Zheng C, Iese Y, Black N, Draper B, Ridley N, Gowing L, Stapinski L, Taye B, Lancaster K, Stjepanović D, Kay-Lambkin F, Jamshidi N, Lubman D, Pastor A, White N, Wilson S, Jaworski AL, Memedovic S, Logge W, Mills K, Seear K, Freeburn B, Lea T, Withall A, Marel C, Boffa J, Roxburgh A, Purcell- Khodr G, Doyle M, Conigrave K, Teesson M, Butler K, Connor J, Morley KC (2021) New Australian guidelines for the treatment of alcohol problems: an overview of recommendations. Med. J. Aust. 215:S3–S32.

22. Hagmann P, Cammoun L, Gigandet X, Meuli R, Honey CJ, Wedeen VJ, Sporns O (2008) Mapping the Structural Core of Human Cerebral Cortex. PLoS Biol. 6:e159.

23. Huang W, King JA, Ursprung WWS, Zheng S, Zhang N, Kennedy DN, Ziedonis D, DiFranza JR (2014) The development and expression of physical nicotine dependence corresponds to structural and functional alterations in the anterior cingulate-precuneus pathway. Brain and Behavior 4:408–417.

24. Hwa L, Besheer J, Kash T (2017) Glutamate plasticity woven through the progression to alcohol use disorder: a multi-circuit perspective. F1000Research 6:298.

25. Kalivas PW, Volkow N, Seamans J (2005) Unmanageable Motivation in Addiction: A Pathology in Prefrontal-Accumbens Glutamate Transmission. Neuron 45:647–650.

26. Kishi T, Miyake N, Okuya M, Sakuma K, Iwata N (2020) N-acetylcysteine as an adjunctive treatment for bipolar depression and major depressive disorder: a systematic review and meta-analysis of double-blind, randomized placebo-controlled trials. Psychopharmacology (Berl.) 237:3481–3487.

27. Krista M, Christopher CL, Peter WK (2003) Prefrontal Glutamate Release into the Core of the Nucleus Accumbens Mediates Cocaine-Induced Reinstatement of Drug-Seeking Behavior. The Journal of Neuroscience 23:3531.

28. Lang PJ, Bradley MM, Cuthbert BN (1997) International affective picture system (IAPS): Technical manual and affective ratings. NIMH Center for the Study of Emotion and Attention 1:3.

29. Li W, van Tol M-J, Li M, Miao W, Jiao Y, Heinze H-J, Bogerts B, He H, Walter M (2014) Regional specificity of sex effects on subcortical volumes across the lifespan in healthy aging. Hum. Brain Mapp. 35:238–247.

30. Logge WB, Morley KC, Haber PS, Baillie AJ (2019) Executive Functioning Moderates Responses to Appetitive Cues: A Study in Severe Alcohol Use Disorder and Alcoholic Liver Disease. Alcohol Alcohol. 54:38–46.

31. Lv H, Wang Z, Tong E, Williams LM, Zaharchuk G, Zeineh M, Goldstein-Piekarski AN, Ball TM, Liao C, Wintermark M (2018) Resting-State Functional MRI: Everything That Nonexperts Have Always Wanted to Know. AJNR Am. J. Neuroradiol. 39:1390–1399.

32. McFarquhar M, McKie S, Emsley R, Suckling J, Elliott R, Williams S (2016) Multivariate and repeated measures (MRM): A new toolbox for dependent and multimodal group-level neuroimaging data. Neuroimage 132:373–389.

33. Morley KC, Baillie A, Van Den Brink W, Chitty KE, Brady K, Back SE, Seth D, Sutherland G, Leggio L, Haber PS (2018) N-acetyl cysteine in the treatment of alcohol use disorder in patients with liver disease: Rationale for further research. Expert Opinion on Investigational Drugs 27:667–675.

34. Morley KC, Lagopoulos J, Logge W, Baillie A, Adams C, Haber PS (2020) Brain GABA levels are reduced in alcoholic liver disease: A proton magnetic resonance spectroscopy study. Addict. Biol. 25:e12702.

35. Morley KC, Perry CJ, Watt J, Hurzeler T, Leggio L, Lawrence AJ, Haber P (2021) New approved and emerging pharmacological approaches to alcohol use disorder: a review of clinical studies. Expert Opin. Pharmacother. 22:1291–1303.

36. Morley KC, Peruch S, Adams C, Towers E, Tremonti C, Watt J, Jamshidi N, Haber PS (2023) N acetylcysteine in the treatment of alcohol use disorder: a randomized, double-blind, placebo-controlled trial. Alcohol Alcohol.:agad044.

37. Nieto-Castanon A, Whitfield-Gabrieli S (2021) CONN functional connectivity toolbox (RRID: SCR_009550), Version 21, in Series CONN functional connectivity toolbox (RRID: SCR_009550), Version 21, Hilbert Press.

38. Pfefferbaum A, Chanraud S, Pitel A-L, Müller-Oehring E, Shankaranarayanan A, Alsop DC, Rohlfing T, Sullivan EV (2011) Cerebral Blood Flow in Posterior Cortical Nodes of the Default Mode Network Decreases with Task Engagement but Remains Higher than in Most Brain Regions. Cereb. Cortex 21:233–244.

39. Prado E, Maes M, Piccoli LG, Baracat M, Barbosa DS, Franco O, Dodd S, Berk M, Vargas Nunes SO (2015) N-acetylcysteine for therapy-resistant tobacco use disorder: a pilot study. Redox Report 20:215–222.

40. Sari Y, Sakai M, Weedman J, Rebec G, Bell R (2015) Ceftriaxone, a Beta-Lactam Antibiotic, Reduces Ethanol Consumption in Alcohol-Preferring Rats. Alcohol Alcohol. 46:239–246.

41. Satterthwaite TD, Elliott MA, Gerraty RT, Ruparel K, Loughead J, Calkins ME, Eickhoff SB, Hakonarson H, Gur RC, Gur RE, Wolf DH (2013) An improved framework for confound regression and filtering for control of motion artifact in the preprocessing of resting-state functional connectivity data. Neuroimage 64:240–256.

42. Schacht JP, Anton RF, Myrick H (2013) Functional neuroimaging studies of alcohol cue reactivity: a quantitative meta-analysis and systematic review. Addict. Biol. 18:121–133.

43. Schaefer A, Kong R, Gordon EM, Laumann TO, Zuo X-N, Holmes AJ, Eickhoff SB, Yeo BTT (2018) Local-Global Parcellation of the Human Cerebral Cortex from Intrinsic Functional Connectivity MRI. Cerebral cortex (New York, N.Y. : 1991) 28:3095-3114.

44. Schulte MHJ, Kaag AM, Boendermaker WJ, van den Brink W, Goudriaan AE, Wiers RW (2019) The effect of N-acetylcysteine and working memory training on neural mechanisms of working memory and cue reactivity in regular cocaine users. Psychiatry Research: Neuroimaging 287:56–59.

45. Schulte MHJ, Wiers RW, Boendermaker WJ, Goudriaan AE, van den Brink W, van Deursen DS, Friese M, Brede E, Waters AJ (2018) The effect of N-acetylcysteine and working memory training on cocaine use, craving and inhibition in regular cocaine users: correspondence of lab assessments and Ecological Momentary Assessment. Addict. Behav. 79:24–31.

46. Skinner HA, Allen BA (1982) Alcohol dependence syndrome: Measurement and validation. J. Abnorm. Psychol. 91:199–209.

47. Sobell LC, Sobell MB (1992) Timeline Follow-Back, in Measuring Alcohol Consumption: Psychosocial and Biochemical Methods, Measuring Alcohol Consumption: Psychosocial and Biochemical Methods (LITTEN RZ, ALLEN JP eds), pp 41–72, Humana Press, Totowa, NJ.

48. Squeglia LM, Baker NL, McClure EA, Tomko RL, Adisetiyo V, Gray KM (2016) Alcohol use during a trial of N-acetylcysteine for adolescent marijuana cessation. Addict. Behav. 63:172–177.

49. Squeglia LM, Tomko RL, Baker NL, McClure EA, Book GA, Gray KM (2018) The effect of N-acetylcysteine on alcohol use during a cannabis cessation trial. Drug Alcohol Depend. 185:17–22.

50. Tapert SF, Cheung EH, Brown GG, Frank LR, Paulus MP, Schweinsburg AD, Meloy MJ, Brown SA (2003) Neural Response to Alcohol Stimuli in Adolescents With Alcohol Use Disorder. Arch. Gen. Psychiatry 60:727–735.

51. Tolomeo S, Yu R (2022) Brain network dysfunctions in addiction: a meta-analysis of resting-state functional connectivity. Translational Psychiatry 12:41.

52. Whitfield-Gabrieli S, Nieto-Castanon A (2012) Conn: A Functional Connectivity Toolbox for Correlated and Anticorrelated Brain Networks. Brain Connect. 2:125–141.

53. Wilcox CE, Abbott CC, Calhoun VD (2019) Alterations in resting-state functional connectivity in substance use disorders and treatment implications. Prog. Neuropsychopharmacol. Biol. Psychiatry 91:79–93.

54. Wrase J, Grüsser SM, Klein S, Diener C, Hermann D, Flor H, Mann K, Braus DF, Heinz A (2002) Development of alcohol-associated cues and cue-induced brain activation in alcoholics. Eur. Psychiatry 17:287–291.

55. Zakiniaeiz Y, Scheinost D, Seo D, Sinha R, Constable RT (2017) Cingulate cortex functional connectivity predicts future relapse in alcohol dependent individuals. NeuroImage: Clinical 13:181–187.

56. Zeng J, Yu S, Cao H, Su Y, Dong Z, Yang X (2021) Neurobiological correlates of cue-reactivity in alcohol-use disorders: A voxel-wise meta-analysis of fMRI studies. Neurosci. Biobehav. Rev. 128:294–310.

