## Supplementary Material for "N-acetyl Cysteine attenuates intrinsic functional connectivity, but not neural alcohol cue reactivity, in treatment-seeking individuals with alcohol use disorder"

### Methods

#### Participants

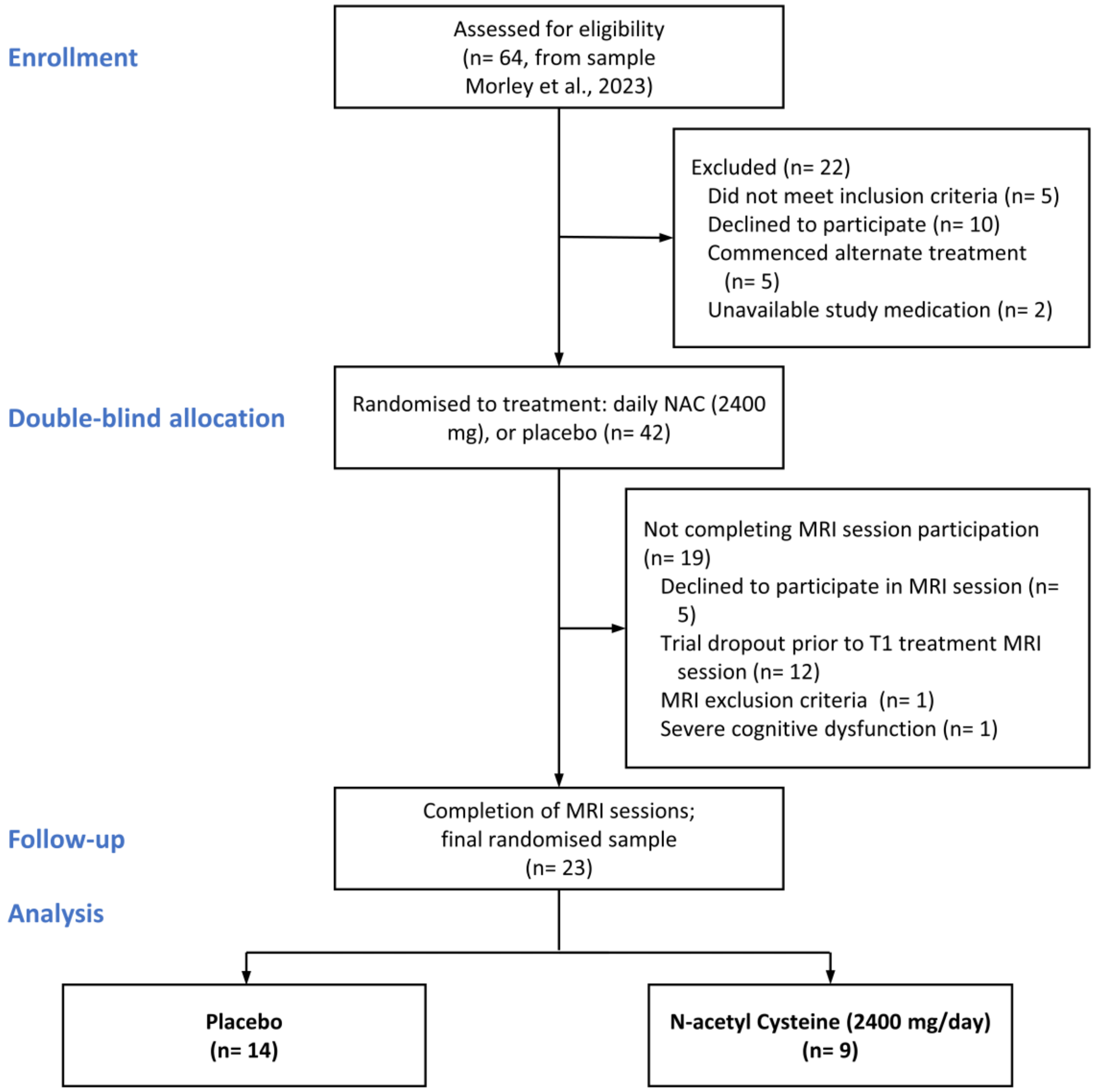


*Supplementary Figure 1 CONSORT flow diagram of participant recruitment and randomization*

### Image processing

*Anatomical data preprocessing*

A total of 2 T1-weighted (T1w) images were found within the input BIDS dataset. All of them were corrected for intensity non-uniformity (INU) with N4BiasFieldCorrection (1), distributed with ANTs 2.3.3 (2), RRID:SCR_004757). The T1w-reference was then skull-stripped with a Nipype implementation of the antsBrainExtraction.sh workflow (from ANTs), using OASIS30ANTs as target template. Brain tissue segmentation of cerebrospinal fluid (CSF), white-matter (WM) and gray-matter (GM) was performed on the brain-extracted T1w using fast (FSL 5.0.9, RRID:SCR_002823, (3). A T1w-reference map was computed after registration of 2 T1w images (after INU-correction) using mri_robust_template (FreeSurfer 6.0.1, (4). Brain surfaces were reconstructed using recon-all (FreeSurfer 6.0.1, RRID:SCR_001847, (5), and the brain mask estimated previously was refined with a custom variation of the method to reconcile ANTs-derived and FreeSurfer-derived segmentations of the cortical gray-matter of Mindboggle (RRID:SCR_002438, (6). Volume-based spatial normalization to two standard spaces (MNI152NLin2009cAsym, MNI152NLin6Asym) was performed through nonlinear registration with antsRegistration (ANTs 2.3.3), using brain-extracted versions of both T1w reference and the T1w template. The following templates were selected for spatial normalization: ICBM 152 Nonlinear Asymmetrical template version 2009c (7), RRID:SCR_008796; TemplateFlow ID: MNI152NLin2009cAsym], FSL's MNI ICBM 152 non-linear 6th Generation Asymmetric Average Brain Stereotaxic Registration Model((8), RRID:SCR_002823; TemplateFlow ID: MNI152NLin6Asym).

*Functional data preprocessing*

For each of the 2 BOLD runs found per subject (across all tasks and sessions), the following preprocessing was performed. First, a reference volume and its skull-stripped version were generated using a custom methodology of fMRIPrep. A B0-nonuniformity map (or fieldmap) was estimated based on two (or more) echo-planar imaging (EPI) references with opposing phase-encoding directions, with 3dQwarp (9) (AFNI 20160207). Based on the estimated susceptibility distortion, a corrected EPI (echo-planar imaging) reference was calculated for a more accurate co-registration with the anatomical reference. The BOLD reference was then co-registered to the T1w reference using bbregister (FreeSurfer) which implements boundary-based registration(10). Co-registration was configured with six degrees of freedom. Head-motion parameters with respect to the BOLD reference (transformation matrices, and six corresponding rotation and translation parameters) are estimated before any spatiotemporal filtering using mcflirt (FSL 5.0.9,(11)). BOLD runs were slice-time corrected using 3dTshift from AFNI 20160207 ((9)x, RRID:SCR_005927). The BOLD time-series (including slice-timing correction when applied) were resampled onto their original, native space by applying a single, composite transform to correct for head-motion and susceptibility distortions. These resampled BOLD time-series will be referred to as preprocessed BOLD in original space, or just preprocessed BOLD. The BOLD time-series were resampled into standard space, generating a preprocessed BOLD run in MNI152NLin2009cAsym space. First, a reference volume and its skull-stripped version were generated using a custom methodology of fMRIPrep. All resamplings can be performed with a single interpolation step by composing all the pertinent transformations (i.e. head-motion transform matrices, susceptibility distortion correction when available, and co-registrations to anatomical and output spaces). Gridded (volumetric) resamplings were performed using antsApplyTransforms (ANTs), configured with Lanczos interpolation to minimize the smoothing effects of other kernels (Lanczos 1964 (12)). Non-gridded (surface) resamplings were performed using mri_vol2surf (FreeSurfer).

*Resting state post-processing of fmriprep outputs*

The eXtensible Connectivity Pipeline (XCP) (13, 14) was used to post-process the outputs of fMRIPrep version 20.2.7 (15, 16), RRID:SCR_016216). XCP was built with Nipype 1.8.5 (Gorgolewski et al. 2011, RRID:SCR_002502). For each of the two BOLD runs found per subject (across all tasks and sessions), the following post-processing was performed. In order to identify high-motion outlier volumes, framewise displacement was calculated using the formula from Power, Mitra (17), with a head radius 40.0 mm. Volumes with framewise displacement greater than 0.4 mm were flagged as high-motion outliers for the sake of later censoring (17). In total, 36 nuisance regressors were selected from the preprocessing confounds, according to the '36P' strategy. These nuisance regressors included six motion parameters, mean global signal, mean white matter signal, mean CSF signal with their temporal derivatives, and the quadratic expansion of six motion parameters, tissues signals and their temporal derivatives (13, 14). Finally, linear trend and intercept terms were added to the regressors prior to denoising. The BOLD data were despiked with 3dDespike. Nuisance regressors were regressed from the BOLD data using linear regression, as implemented in nilearn 0.10.0 (18). Any volumes censored earlier in the workflow were then interpolated in the residual time series produced by the regression. The interpolated timeseries were then band-pass filtered using a(n) second-order Butterworth filter, in order to retain signals within the 0.01-0.08 Hz frequency band. The filtered, interpolated time series were then re-censored to remove high-motion outlier volumes. The denoised BOLD was smoothed using Nilearn with a Gaussian kernel (FWHM=6.0 mm).

Processed functional timeseries were extracted from the residual BOLD signal with Nilearn's (version 0.10.0, (18)) NiftiLabelsMasker was used for the Schaefer 17-network 400 parcel atlas (19). Corresponding pair-wise functional connectivity between all regions was computed for each atlas, which was operationalized as the Pearson's correlation of each parcel's unsmoothed timeseries. In cases of partial coverage, uncovered voxels (values of all zeros or NaNs) were either ignored, when the parcel had >50.0% coverage, or were set to zero, when the parcel had <50.0% coverage.

Many internal operations of XCP use AFNI (9, 20), ANTS (21), TemplateFlow version 0.8.1 (22), matplotlib version 3.4.3 (23), Nibabel version 5.0.1 (Brett et al. 2022), Nilearn version 0.10.0 (18), numpy version 1.22.4 (24), pybids version 0.15.5 (Yarkoni et al. 2019) (25), and scipy version 1.9.1(26). For more details, see the xcp_d website https://xcp-d.readthedocs.io.

### Supplementary Results

*Supplementary Table 1 Linear Mixed Effects models for AUQ*

| | T0 (Baseline) | | | T1 (During Treatment) | | | T0 vs T1 Full model | | |
|---|---|---|---|---|---|---|---|---|---|
| Predictors | β | CI | p | β | CI | p | β | CI | p |
| (Intercept) | 31.89 | 13.89 – 49.88 | **.001** | 33.84 | 18.80 – 48.88 | **<.001** | 29.86 | 14.90 – 44.82 | **<.001** |
| Treatment (NAC) | -3.52 | -12.18 – 5.13 | .425 | -5.97 | -13.21 – 1.26 | .105 | -2.23 | -10.49 – 6.02 | .596 |
| Pre/Post Scan (Post) | 5.27 | -2.14 – 12.68 | .164 | -5.52 | -17.10 – 6.07 | .35 | 1.26 | -12.70 – 15.22 | .86 |
| Age | -0.28 | -0.64 – 0.07 | .12 | -0.33 | -0.63 – -0.03 | **.029** | -0.24 | -0.53 – 0.05 | .104 |
| Treatment (NAC) * Pre/Post Scan (Post) | 0.19 | -3.37 – 3.75 | .916 | -0.63 | -6.20 – 4.94 | .825 | -0.64 | -9.56 – 8.28 | .888 |
| Pre/Post Scan * age | -0.09 | -0.24 – 0.05 | .216 | 0.12 | -0.11 – 0.35 | .311 | -0.01 | -0.28 – 0.26 | .942 |
| Session (T1) | | | | | | | -0.43 | -5.98 – 5.12 | .88 |
| Session (T1) * Treatment (NAC) | | | | | | | -0.68 | -9.55 – 8.19 | .88 |
| Session * Pre/Post Scan | | | | | | | -0.57 | -8.42 – 7.28 | .887 |
| Session * Treatment * Pre/Post Scan | | | | | | | -0.65 | -13.20 – 11.90 | .919 |
| **Random Effects** | | | | | | | | | |
| $\sigma^2$ | 8.32 | | | 20.35 | | | 56.13 | | |
| $\tau_{00}$ | 89.87 ID | | | 48.29 ID | | | 39.49 ID | | |
| ICC | 0.92 | | | 0.7 | | | 0.41 | | |
| N | 22 ID | | | 22 ID | | | 23 ID | | |
| Observations | 44 | | | 44 | | | 92 | | |
| Marginal $R^2$ / Conditional $R^2$ | 0.167 / 0.929 | | | 0.245 / 0.776 | | | 0.115 / 0.480 | | |

Note. Reference category for predictor shown in brackets. $\sigma^2$ = random effects variance; $\tau_{00}$ = random intercept variance ICC = intra-class correlation coefficient; ID = individual participants (random factor).

**Cue reactivity Exploratory Whole Brain Analyses**

Across the whole sample, irrespective of treatment group, the Alcohol images elicited increased BOLD activation compared to the Control images during fMRI cue reactivity. This was seen in 5 clusters, including one encompassing the left parahippocampal gyrus and fusiform gyrus ($P_{FWE\text{-}corr} < .030$, 280 voxels) and one cluster including the right parahippcampal and fusiform gyrus ($P_{FWE\text{-}corr} < .014$, 454 voxels). Two clusters were observed within the occipital cortex and adjacent regions, primarily the left supramarginal gyrus, angular gyrus, and middle temporal gyrus($P_{FWE\text{-}corr} < .023$, 318 voxels),  the right angular gyrus and supramarginal gyrus ($P_{FWE\text{-}corr} < .038$, 237 voxels). One cluster encompassed the posterior cingulate and left precuneus  ($P_{FWE\text{-}corr} < .030$, 277 voxels).

No other main effects of time or treatment group were found, and no main effects of covariates. No two-way interactions were seen between condition, time, treatment group were seen. There was a significant three-way interaction of condition, time, and antidepressant use in a cluster that spanned the bilateral thalamus, and right medial dorsal nucleus of the thalamus, extra-nuclear,  and extending into the left middle and superior temporal gyri and left insula ($P_{FWE\text{-}corr} < .013$, 682 voxels).

**ALC > CON contrasts at T0**

Results are presented in Supplementary Table 2. There were no significant effects observed at T0 for any of the 5 ROIs according to any of the variables including treatment group, drinks per drinking day, age, ArLD, or antidepressant use, or any two-way interactions (*p*'s > .0413), indicating that there were no differences in cue reactivity at baseline according to treatment group.

**ALC > CON contrasts at T1**

Results are presented in Supplementary Table 2. At T1 no significant main treatment effect seen for the ROIs. There was a significant effect for covariates, with a significant main effect of presence of alcoholic liver disease for the right DLPFC, with those with ArLD showing increased alcohol cue reactivity overall ($p$ =.024). There was also a main effect of antidepressant use with those on antidepressants showing increased alcohol cue reactivity ($p$ = .013).

*Supplementary Table 2 ANCOVAs for ALC > Con contrast across 5 ROIs during T0 and T1*

| Predictors | Bl VMPFC | L Caudate Body | R Caudate Body | L DLPFC | R DLPFC |
|---|---|---|---|---|---|
| T0 | | | | | |
| Treatment | F(1,15)=1.2, p = .292 | F(1,15)=0.15, p = .704 | F(1,15)=0.13, p = .724 | F(1,15)=0.02, p = .878 | F(1,15)=0.04, p = .841 |
| Age | F(1,15)=2.59, p = .129 | F(1,15)=0.22, p = .646 | F(1,15)=0.25, p = .627 | F(1,15)=0.09, p = .769 | F(1,15)=1.38, p = .258 |
| Alcoholic Liver Disease (Yes) | F(1,15)=3.66, p = .075 | F(1,15)=0.1, p = .762 | F(1,15)=0.12, p = .737 | F(1,15)=0.44, p = .518 | F(1,15)=0.4, p = .536 |
| Antidepressant Use (Yes) | F(1,15)=0, p = .99 | F(1,15)=0.53, p = .477 | F(1,15)=0.91, p = .356 | F(1,15)=0.14, p = .718 | F(1,15)=1.56, p = .231 |
| Drinks per drinking day | F(1,15)=1.08, p = .315 | F(1,15)=0.01, p = .922 | F(1,15)=0.21, p = .657 | F(1,15)=0.18, p = .674 | F(1,15)=0.01, p = .913 |
| Treatment * Age | F(1,15)=0.02, p = .884 | F(1,15)=0.19, p = .668 | F(1,15)=0.3, p = .595 | F(1,15)=0, p = .991 | F(1,15)=0.04, p = .844 |
| T1 | | | | | |
| Treatment | F(1,14)=0.27, p = .614 | F(1,14)=1.6, p = .227 | F(1,14)=0.08, p = .776 | F(1,14)=2.01, p = .178 | F(1,14)=0.18, p = .682 |
| Age | F(1,14)=0, p = .954 | F(1,14)=0, p = .971 | F(1,14)=0, p = .99 | F(1,14)=4.54, p = .051 | F(1,14)=6.04, p = **.028** |
| Alcoholic Liver Disease (Yes) | F(1,14)=2.25, p = .156 | F(1,14)=0.05, p = .834 | F(1,14)=0, p = .983 | F(1,14)=2.96, p = .107 | F(1,14)=1.1, p = .312 |
| Antidepressant Use (Yes) | F(1,14)=0.03, p = .865 | F(1,14)=0.79, p = .389 | F(1,14)=0.19, p = .667 | F(1,14)=0, p = .979 | F(1,14)=0.39, p = .541 |
| Drinks per drinking day | F(1,14)=2.46, p = .139 | F(1,14)=8.73, p = **.011** | F(1,14)=1.74, p = .208 | F(1,14)=4.59, p = .05 | F(1,14)=0, p = .959 |
| Treatment Days | F(1,14)=1.78, p = .203 | F(1,14)=4.78, p = .046 | F(1,14)=1.36, p = .263 | F(1,14)=3.8, p = .072 | F(1,14)=1.98, p = .182 |
| Treatment * Age | F(1,14)=0.87, p = .366 | F(1,14)=0.42, p = .527 | F(1,14)=0.36, p = .559 | F(1,14)=2.89, p = .111 | F(1,14)=0.57, p = .462 |

*Supplementary Table 3 Linear Mixed Models for ALC vs CON contrast across 5 ROIs*

| | Bl VMPFC | | | L Caudate | | | R Caudate | | | L DLPFC | | | R DLPFC | | |
|---|---|---|---|---|---|---|---|---|---|---|---|---|---|---|---|
| Predictors | β | CI | p | β | CI | p | β | CI | p | β | CI | p | β | CI | p |
| (Intercept) | 0.22 | -0.20 – 0.64 | .296 | -0.12 | -0.52 – 0.27 | .534 | -0.06 | -0.45 – 0.32 | .747 | -0.04 | -0.38 – 0.29 | .803 | -0.11 | -0.56 – 0.34 | .623 |
| Session | -0.14 | -0.36 – 0.09 | .227 | 0 | -0.21 – 0.21 | .999 | -0.01 | -0.22 – 0.19 | .905 | -0.07 | -0.24 – 0.10 | .421 | -0.12 | -0.33 – 0.08 | .236 |
| Treatment | -0.06 | -0.33 – 0.20 | .631 | 0.1 | -0.15 – 0.34 | .43 | 0.08 | -0.17 – 0.32 | .531 | 0.02 | -0.19 – 0.22 | .878 | 0 | -0.26 – 0.27 | .982 |
| Age | 0 | -0.01 – 0.01 | .906 | 0 | -0.01 – 0.01 | .724 | 0 | -0.01 – 0.01 | .796 | 0 | -0.01 – 0.01 | .974 | 0 | -0.01 – 0.01 | .782 |
| Antidepressant Use | -0.13 | -0.33 – 0.06 | .177 | -0.09 | -0.28 – 0.09 | .321 | -0.07 | -0.25 – 0.11 | .451 | -0.05 | -0.21 – 0.10 | .501 | -0.05 | -0.26 – 0.16 | .656 |
| Alcoholic Liver Disease | 0.11 | -0.09 – 0.31 | .278 | 0.14 | -0.05 – 0.33 | .137 | 0.09 | -0.09 – 0.27 | .334 | 0.14 | -0.02 – 0.30 | .086 | 0.17 | -0.05 – 0.38 | .129 |
| Session * Treatment | 0.19 | -0.17 – 0.54 | .3 | 0.04 | -0.29 – 0.38 | .796 | -0.02 | -0.34 – 0.31 | .912 | 0.12 | -0.15 – 0.40 | .364 | 0.07 | -0.25 – 0.40 | .651 |
| **Random Effects** | | | | | | | | | | | | | | | |
| $\sigma^2$ | 0.09 | | | 0.08 | | | 0.07 | | | 0.05 | | | 0.07 | | |
| $\tau_{00}$ | 0.00 ID | | | 0.00 ID | | | 0.00 ID | | | 0.00 ID | | | 0.01 ID | | |
| ICC | | | | | | | | | | 0.03 | | | 0.16 | | |
| N | 23 ID | | | 23 ID | | | 23 ID | | | 23 ID | | | 23 ID | | |
| Observations | 46 | | | 46 | | | 46 | | | 46 | | | 46 | | |
| Marginal $R^2$ | 0.084 | | | 0.079 | | | 0.035 | | | 0.093 | | | 0.085 | | |

P-threshold < .029 (Bonferroni's adjustment corrected)

Note. Reference category for predictor shown in brackets. $\sigma^2$ = random effects variance; $\tau_{00}$ = random intercept variance ICC = intra-class correlation coefficient; ID = individual participants (random factor); Bl = bilateral, L = left, R = right, DLPFC = dorsolateral prefrontal cortex.

*Supplementary Table 4 Connections with significant seed region posterior cingulate 9 from Schaeffer-400 atlas parcellation*

| Seed ROI | Connection Side | Connection target | Test statistic | p-unc | p-FDR |
|---|---|---|---|---|---|
| Dorsal Attentional Posterior Cingulate 9 | | | F(2,17)=20.75 | 0.000027 | .010 |
| | Right | Somatomotor A 6 | T(18)=-6.13 | 0.000009 | .003 |
| | Right | Somatomotor A 10 | T(18)=-5.06 | 0.000081 | .013 |
| | Left | Somatomotor A 7 | T(18)=-4.95 | 0.000104 | .013 |
| | Left | Somatomotor A 10 | T(18)=-4.76 | 0.000156 | .015 |
| | Left | Visual B Striate Calcarine 1 | T(18)=-4.52 | 0.000266 | .020 |
| | Left | Somatomotor B Central 5 | T(18)=-4.42 | 0.000332 | .021 |
| | Right | Somatomotor A 2 | T(18)=-4.11 | 0.000657 | .035 |
| | Right | Somatomotor A 7 | T(18)=-4.04 | 0.000765 | .036 |
| | Right | Somatomotor A 4 | T(18)=-3.63 | .002 | .081 |
| | Right | Temporal Parietal 2 | T(18)=-3.51 | .002 | .093 |
| | Right | Somatomotor A 11 | T(18)=-3.47 | .003 | .094 |
| | Right | Salience/Ventral Attention B Medial Posterior PFC 1 | T(18)=-3.37 | .003 | .106 |
| | Right | Visual B Extra-striate Superior 2 | T(18)=-3.3 | .004 | .116 |
| | Left | Somatomotor A 3 | T(18)=-3.21 | .005 | .130 |
| | Right | Visual A Extra-striate Inferior 10 | T(18)=-3.16 | .005 | .131 |
| | Left | Somatomotor A 13 | T(18)=-3.15 | .006 | .131 |
| | Right | Somatomotor A 3 | T(18)=-2.98 | .008 | .161 |
| | Left | Somatomotor B Central 4 | T(18)=-2.94 | .009 | .161 |

| | | | | | |
|---|---|---|---|---|---|
| | Left | Temporal Parietal 3 | T(18)=-2.94 | .009 | .161 |
| | Left | Visual B Striate Calcarine 2 | T(18)=-2.93 | .009 | .161 |
| | Right | Visual B Striate Calcarinel 1 | T(18)=-2.91 | .009 | .161 |
| | Left | Visual A Extrastriate 10 | T(18)=-2.91 | .009 | .161 |

### References

1. Tustison NJ, Avants BB, Cook PA, Zheng Y, Egan A, Yushkevich PA, et al. N4ITK: Improved N3 Bias Correction. IEEE Trans Med Imaging. 2010;29(6):1310-20.

2. Avants BB, Epstein CL, Grossman M, Gee JC. Symmetric diffeomorphic image registration with cross-correlation: Evaluating automated labeling of elderly and neurodegenerative brain. Med Image Anal. 2008;12(1):26-41.

3. Zhang Y, Brady M, Smith S. Segmentation of brain MR images through a hidden Markov random field model and the expectation-maximization algorithm. IEEE Trans Med Imaging. 2001;20(1):45-57.

4. Reuter M, Rosas HD, Fischl B. Highly accurate inverse consistent registration: A robust approach. Neuroimage. 2010;53(4):1181-96.

5. Dale AM, Fischl B, Sereno MI. Cortical Surface-Based Analysis: I. Segmentation and Surface Reconstruction. Neuroimage. 1999;9(2):179-94.

6. Klein A, Ghosh SS, Bao FS, Giard J, Häme Y, Stavsky E, et al. Mindboggling morphometry of human brains. PLoS Comput Biol. 2017;13(2):e1005350.

7. Fonov VS, Evans AC, McKinstry RC, Almli CR, Collins DL. Unbiased nonlinear average age-appropriate brain templates from birth to adulthood. Neuroimage. 2009;47:S102.

8. Evans AC, Janke AL, Collins DL, Baillet S. Brain templates and atlases. Neuroimage. 2012;62(2):911-22.

9. Cox RW, Hyde JS. Software tools for analysis and visualization of fMRI data. NMR Biomed. 1997;10(4-5):171-8.

10. Greve DN, Fischl B. Accurate and robust brain image alignment using boundary-based registration. Neuroimage. 2009;48(1):63-72.

11. Jenkinson M, Bannister P, Brady M, Smith S. Improved Optimization for the Robust and Accurate Linear Registration and Motion Correction of Brain Images. Neuroimage. 2002;17(2):825-41.

12. Lanczos C. Evaluation of Noisy Data. Journal of the Society for Industrial and Applied Mathematics Series B Numerical Analysis. 1964;1(1):76-85.

13. Ciric R, Wolf DH, Power JD, Roalf DR, Baum GL, Ruparel K, et al. Benchmarking of participant-level confound regression strategies for the control of motion artifact in studies of functional connectivity. Neuroimage. 2017;154:174-87.

14. Satterthwaite TD, Elliott MA, Gerraty RT, Ruparel K, Loughead J, Calkins ME, et al. An improved framework for confound regression and filtering for control of motion artifact in the preprocessing of resting-state functional connectivity data. Neuroimage. 2013;64:240-56.

15. Esteban O, Ciric R, Finc K, Blair RW, Markiewicz CJ, Moodie CA, et al. Analysis of task-based functional MRI data preprocessed with fMRIPrep. Nat Protoc. 2020;15(7):2186-202.

16. Esteban O, Markiewicz CJ, Blair RW, Moodie CA, Isik AI, Erramuzpe A, et al. fMRIPrep: a robust preprocessing pipeline for functional MRI. Nature Methods. 2019;16(1):111-6.

17. Power JD, Mitra A, Laumann TO, Snyder AZ, Schlaggar BL, Petersen SE. Methods to detect, characterize, and remove motion artifact in resting state fMRI. Neuroimage. 2014;84:320-41.

18. Abraham A, Pedregosa F, Eickenberg M, Gervais P, Mueller A, Kossaifi J, et al. Machine learning for neuroimaging with scikit-learn. Front Neuroinform. 2014;8.

19. Schaefer A, Kong R, Gordon EM, Laumann TO, Zuo X-N, Holmes AJ, et al. Local-Global Parcellation of the Human Cerebral Cortex from Intrinsic Functional Connectivity MRI. Cerebral cortex (New York, NY : 1991). 2018;28(9):3095-114.

20. Cox RW. AFNI: Software for Analysis and Visualization of Functional Magnetic Resonance Neuroimages. Comput Biomed Res. 1996;29(3):162-73.

21. Avants BB, Tustison N, Song G. Advanced normalization tools (ANTS). Insight j. 2009;2(365):1-35.

22. Ciric R, Thomas AW, Esteban O, Poldrack RA. Differentiable programming for functional connectomics. arXiv preprint arXiv:220600649. 2022.

23. Hunter JD. Matplotlib: A 2D Graphics Environment. Computing in Science & Engineering. 2007;9(03):90-5.

24. Harris CR, Millman KJ, van der Walt SJ, Gommers R, Virtanen P, Cournapeau D, et al. Array programming with NumPy. Nature. 2020;585(7825):357-62.

25. Yarkoni T, Markiewicz CJ, de la Vega A, Gorgolewski KJ, Salo T, Halchenko YO, et al. PyBIDS: Python tools for BIDS datasets. J Open Source Softw. 2019;4(40).

26. Virtanen P, Gommers R, Oliphant TE, Haberland M, Reddy T, Cournapeau D, et al. SciPy 1.0: fundamental algorithms for scientific computing in Python. Nature Methods. 2020;17(3):261-72.
